# Novel *in vivo* measurement of muscle total carnitine concentration reveals potential mechanism linking mitochondrial dysfunction and lipid accumulation

**DOI:** 10.64898/2026.07.28.26359098

**Authors:** Katherine Schon, Laura Watson, Heather Biggs, Kritika Grover, Ajay Thankamony, Emma Harrison, Michele Ferraro, Jelle van den Ameele, Chris Boesch, Jun Han, David Schibli, Albert Koulman, Krishna Chatterjee, Rita Horvath, Graham Kemp, Patrick Chinnery, Alison Sleigh

**Affiliations:** Department of Clinical Neurosciences, University of Cambridge, Cambridge Biomedical Campus, Cambridge, UK; Medical Research Council Mitochondrial Biology Unit, University of Cambridge, Cambridge, UK; Department of Genomic Medicine, University of Cambridge, Cambridge Biomedical Campus, Cambridge, UK; National Institute for Health and Care Research Cambridge Clinical Research Facility, Cambridge University Hospitals NHS Foundation Trust, Cambridge Biomedical Campus, Cambridge, UK; Institute of Metabolic Science Metabolic Research Laboratories, University of Cambridge, Cambridge Biomedical Campus, Cambridge, UK; Department of Paediatrics, University of Cambridge, Cambridge Biomedical Campus, Cambridge, UK; University Bern, Bern, Switzerland; UVic Genome BC Proteomics Centre, University of Victoria, Victoria, Canada; Department of Musculoskeletal & Ageing Science, University of Liverpool, Liverpool, UK

## Abstract

Free carnitine is essential to mitochondrial health by buffering the free acetyl-CoA pool and thereby maintaining energy production. It is also responsible for transporting long-chain fatty acids into the mitochondria for oxidation. Almost all the body’s carnitine is in muscle, and plasma concentrations do not reflect tissue content, but there are as yet no non-invasive techniques to assess muscle total or free carnitine. Here we describe a novel non-invasive postprocessing method, using standard ^1^H magnetic resonance spectroscopy data, for quantifying muscle total and free carnitine concentrations, which includes an orientation-visibility and spectral fitting component, and consideration of interfering metabolites. We demonstrate the importance of the orientation correction even within one muscle group (accounting for up to 1.9-fold difference within one muscle group and 2.9-fold difference in signal between muscles), show its good reproducibility (CoV 8-12%), and validate the results with mass spectrometry measurements in muscle biopsy samples. We apply this method in a group of patients with genetic mitochondrial disease, to investigate the relationship between mitochondrial dysfunction and muscle lipid accumulation. As predicted muscle total and free carnitine were lower in patients with disease and correlated with the degree of mitochondrial dysfunction and lipid accumulation. Further, robust spatial correlations of total carnitine and muscle lipid imply heterogeneity in mitochondrial function. Our findings suggest that increasing muscle carnitine stores could ameliorate the metabolic effects of and disorders related to mitochondrial dysfunction. Furthermore, it has not usually been known in supplementation studies whether l-carnitine actually reached the target tissue. We suggest that this novel method has significant potential for informing on physiology and pathophysiology, and as a biomarker in monitoring treatment response, investigative drug discovery, and personalised medicine.

## Main

Mitochondrial dysfunction plays an increasingly recognised role in a range of pathologies of clinical and public health importance: examples include frailty, sarcopenia, neurodegeneration, obesity, hepatic steatosis and cardiovascular disease. Free carnitine has a number of relevant metabolic roles relevant to mitochondrial function ^1^. In addition to its well-known role in transporting long-chain fatty acids into mitochondria for oxidation, it helps maintain mitochondrial health by buffering the free acetyl-CoA pool, particularly during exercise, thus reducing accumulation of toxic acyl-CoA compounds and maintaining energy production. It is also thought to protect against oxidative stress and have anti-inflammatory properties and is involved in metabolic pathways of protein balance and cell volume. L-carnitine supplementation is life-saving in primary carnitine deficiency, and in a number of other situations supplementation with either L-carnitine or acetyl-L-carnitine has beneficial effects ^1^: these include complex I deficiency ^2^ and mitochondrial fatty acid oxidation disorders ^3^, cardiac disease, neurodegeneration, liver disease, kidney disease, obesity ^4^ and diabetes, aging, and athletic performance. However, it has sometimes failed to show beneficial effects and this, along with uncertainty of potential detrimental effects of increasing trimethylamine N-oxide ^5^, has perhaps reduced enthusiasm for research into carnitine supplementation.

A significant limitation of these studies is uncertainty over how much the supplement reached the target tissues. As > 95 % of total body carnitine is in skeletal muscle and only 0.5 % in plasma, plasma concentrations cannot be taken as reliable measures of cellular concentrations ^6–8^. Biopsy sampling is invasive, unable to sample the same region twice, and being highly localised may also be unrepresentative of the wider anatomical distribution. In principle, MRS is an ideal way to non-invasively make localised, quantifiable and reproducible measurements of tissue carnitine concentrations. Total carnitine is largely composed of free carnitine (80 - 90 %), with the remainder being acetylcarnitine, and to a lesser extent other acylcarnitines. Free carnitine plays a crucial role in buffering the mitochondrial acetyl-CoA pool, converting it to acetylcarnitine and the acetylcarnitine to carnitine ratio is thought to relate to mitochondrial acetyl-CoA to CoA ratio ^9^. Although acetylcarnitine can be measured using ^1^H-MRS ^10^, measurement of free and total carnitine poses a number of technical problems which are avoided or mitigated in the new method we describe here.

The muscle ^1^H MRS trimethylamine (TMA) resonance has been assumed to be carnitine by Ren et al. ^11,12^, and suggested to be predominantly carnitine by another group ^13^. However, interpretation of this resonance is complicated: firstly, it is subject to orientation-dependent NMR visibility effects ^10,14,15^; secondly, other metabolites can appear under the TMA resonance. Here we address these issues by allowing for orientation in spectral fitting and concentration calculation, and systematically considering potentially ‘contaminating’ metabolites. The method very conveniently uses features that are inherent within the ^1^H-MRS spectrum: namely utilising the creatine CH_2_ resonance for assessment of muscle fibre orientation relative to the main magnetic field, and other peak resonances for contribution of other metabolites.

The method we describe measures muscle total carnitine concentration at 3T quickly and with good reproducibility. We demonstrate the importance of the orientation correction and validate the results with mass spectrometry measurements in muscle biopsy samples in a subset of participants. We suggest that it has significant potential for informing on physiology and pathophysiology, and as a biomarker in monitoring treatment response, investigative drug discovery, and personalised medicine. As an example of its application to clinical research we use it in a group of patients with genetic mitochondrial disease, to throw light on the poorly-understood mechanism of the association of mitochondrial dysfunction with skeletal muscle lipid accumulation ^16^. As a respiratory chain or fatty acid oxidation defect is known to cause accumulation of acylcarnitines, which are exported from the cell ^17^, we hypothesised that this would lower the total muscle carnitine content in mitochondrial disease.

## Results

### Development of a method to measure skeletal muscle total carnitine concentration *in vivo*

#### Correcting for muscle fibre orientation is necessary

In order to compare the variation in muscle fibre orientation relative to the main magnetic field (B_o_) between people and muscle groups, we studied the splitting of the creatine CH_2_ (Cr2) resonance in healthy individuals (see Dataset 1 within Methods). Example spectra acquired at 3T and fit for the soleus (SOL) and tibialis anterior (TA) muscles are shown in Fig 1A. Fig. 1B shows the inter-subject variation in splitting between the creatine CH_2_ (Cr2) resonances, which reflects variation in fibre orientation relative to B_o_: this differed as expected between muscles, but also within the same muscle despite standardised leg and foot positioning. This translates into significant inter-subject differences in NMR visibility, shown in Fig. 1B by the hypothetical signal loss of the TMA and methyl of creatine (Cr3) resonances: the expected range of TMA signal loss was for SOL (0-40%), VL (9-53%) and TA (42-66%); the Cr3 resonance, less influenced by NMR visibility effects, it was for SOL (0-21%), VL (5-28%) and TA (22-35%).

**Fig. 1.**
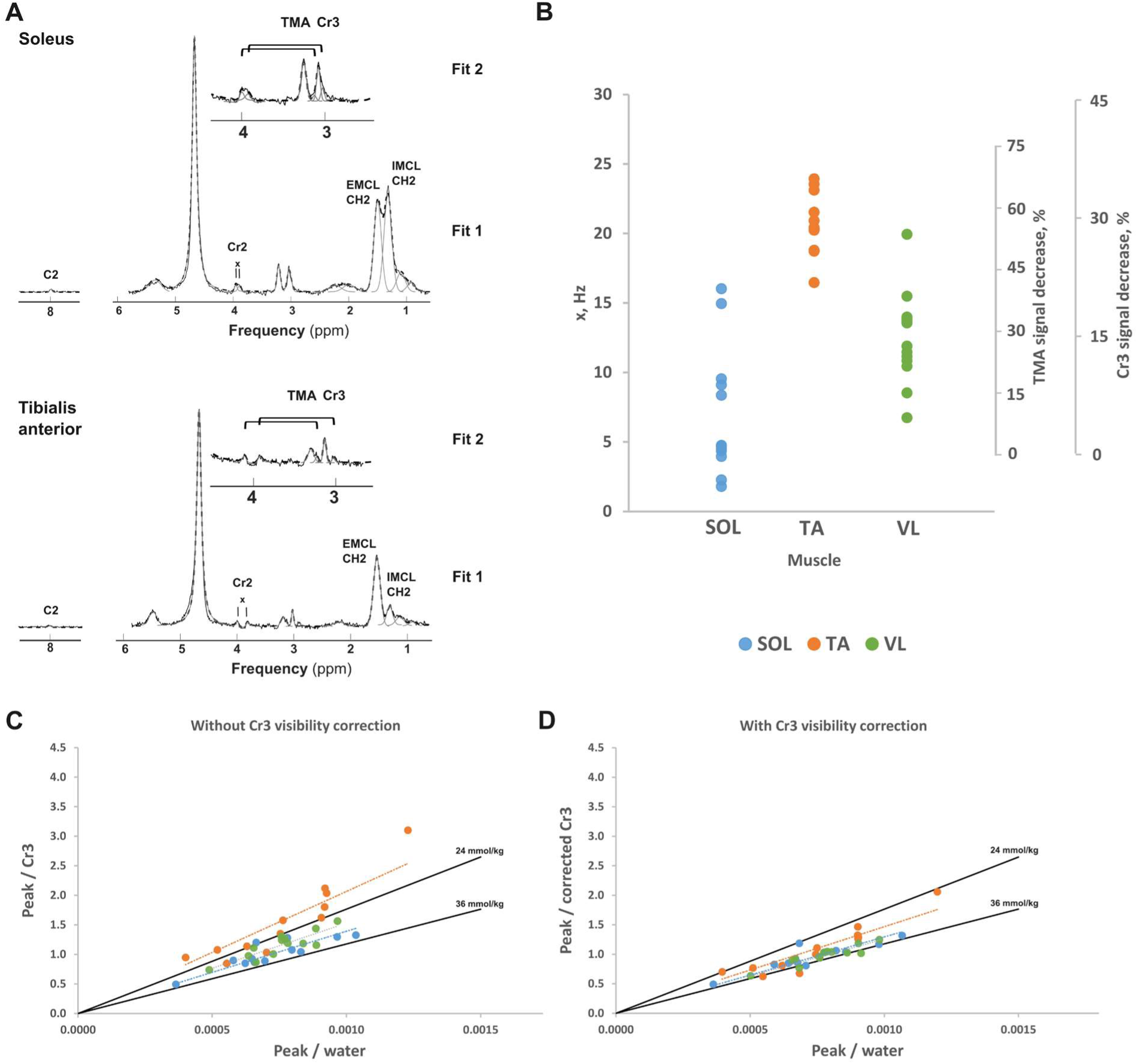
Validation of orientation-dependent visibility correction. Panel (a) shows a representative soleus and tibialis anterior spectra, acquired and fitted using routine 1 (upper insert routine 2). Routine 1 obtained measures of carnosine C2, extramyocellular (EMCL) and intramyocellular lipids (IMCL), as well as frequency splitting of Cr2 resonances (x). Routine 2 fitted the Cr3 resonance as a triplet, with outer coupled resonances constrained in frequency relative to the fitted Cr2 resonances (Supplementary Table 1), and generated values for the total Cr3 and TMA. Solid lines represent original spectra, dotted the estimated fit, and individual resonances in grey. Panel (b) shows the splitting separation of Cr2 resonances in dataset 1 for soleus (SOL), tibialis anterior (TA) and vastus lateralis (VL); the right-hand y-axis shows the orientation-dependent expected signal decreases of carnitine and Cr3. Panels (c) & (d) show the validation of the NMR visibility correction on the Cr3 resonance using dataset 1 by comparing a peak within the spectrum (the corrected TMA peak) relative to Cr3 (y-axis) and relative to water (x-axis) without (c) and with (d) visibility correction of the Cr3 peak. The solid black lines show the reported range of biopsy creatine concentrations (24 – 36 mmol/kg ww); also shown are the colour-coded regression lines through the origin for each muscle.

Making the presumption (standard in quantitative MRS work) that intersubject variation in water and total creatine concentration in human skeletal muscle can to a first approximation be ignored, we tested the performance of the orientation-dependent correction on the Cr3 resonance by comparing the water/creatine calibration conversion (Fig. 1 C&D). Correction yielded creatine concentrations within the literature-derived range of 24-36 mmol/kg wet weight (ww). Regressing through the origin for each muscle yielded total creatine concentrations for SOL, VL and TA respectively of 32.7, 33.6 and 28.9 mmol/kg ww with the visibility correction, compared to 30.6, 26.6 and 18.7 mmol/kg ww without visibility correction. The gradient of the regression lines through the origin for each muscle (Fig. 1D) yield conversion factors, k = 1295, 1468, and 1260 for SOL, TA and VL respectively, for conversion to relative water when no non-water suppressed data were acquired (Dataset 2).

### Analytical specificity

Table 1 lists metabolites that could, in principle, contribute to the TMA resonance. *Carnosine* is likely the most important in quantitative terms, and we corrected for this by calculating total carnitine concentration as the corrected TMA signal minus the carnosine contribution expected at the TMA frequency, calculated from the corrected carnosine signal at ∼ 8.0 ppm (see Methods).

**Table 1.**
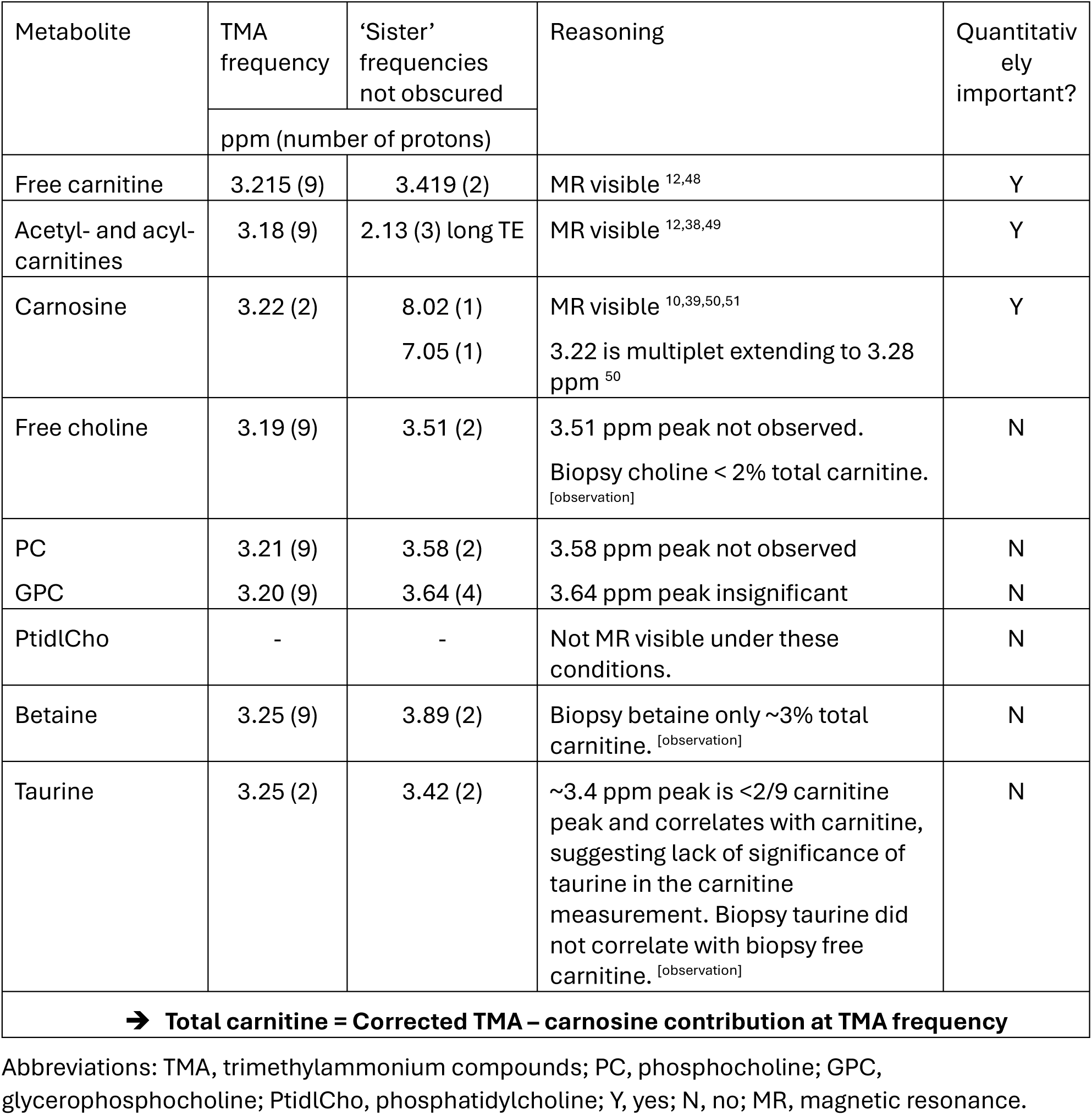
Metabolites potentially contributing to the TMA resonance.

We then assessed the contribution of *taurine* to the TMA resonance by using this corrected carnitine concentration to compare to the visibility and T2 corrected signal at ∼3.4 ppm (S_o 3.4_) that in theory includes contributions from both free carnitine and taurine. In healthy volunteers S_o 3.4_ correlated with the measured TMA-carnitine concentration (Supplementary Fig. S2; r_s_=0.594, 0.810, 0.747 and p=0.042, 0.001, 0.003 for SOL, TA, VL respectively) and was, on average, less than 2/9 S_o CTN_ (relative proton density of carnitine at 3.4 & 3.2 ppm), ruling out taurine as a significant contributor to the TMA-carnitine measurement. In the 8 patients (dataset 3 within Methods), the 3.4 ppm resonance(s) were only detectable in 3 participants from the SOL and TA and 4 in the VL, and all but one were lower than the expected 2/9 S_o CTN_. There was no correlation of biopsy taurine with biopsy free or total carnitine (p=0.6).

Fig. 2 shows that the MRS measures of total carnitine are in good agreement with biopsy measurement: the absolute agreement intraclass correlation coefficient (ICC) was 0.905, p = 0.01.

**Fig. 2.**
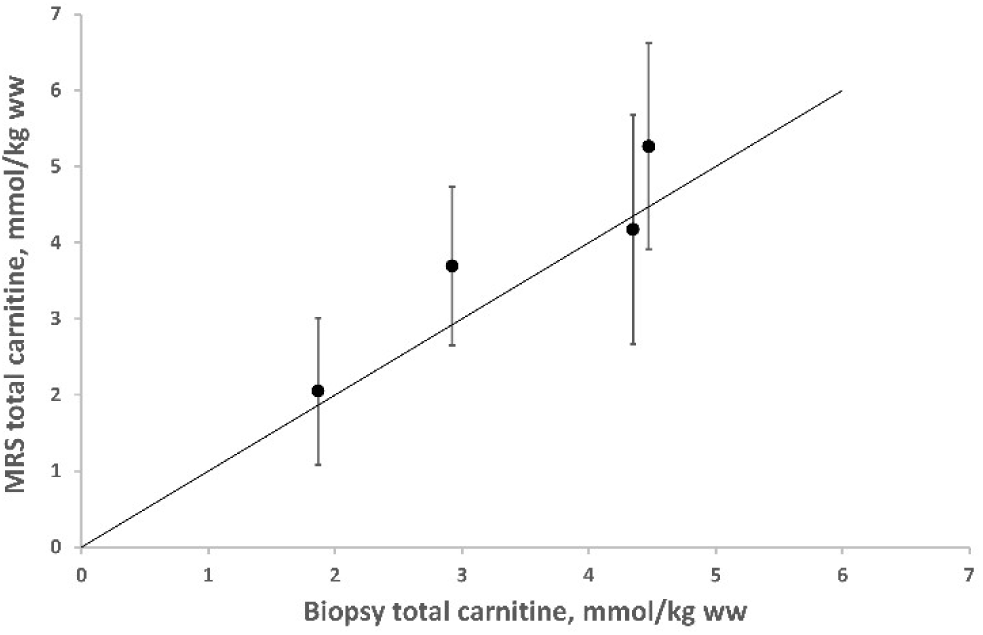
Validation of ^1^H MRS measured carnitine concentration. Validation against biopsy carnitine of MRS total carnitine measured as corrected TMA peak minus the expected carnosine concentration at this frequency, in 4 patients with mitochondrial disease. The MRS carnitine concentrations are the average over the three muscles, and error bars represent standard errors. The line of identity is the solid black line.

#### Total carnitine measurements are reproducible

Reproducibility of total carnitine concentrations was assessed using data from a study of healthy males who undertook two MRS exams 20 h apart, at 8 h and 28 h of a fasting intervention. This is a pragmatic measure of CoV and must be regarded as an upper limit: little is known about carnitine changes in fasting, although it is thought to turn over only slowly. The average carnitine reproducibility CoV for SOL and TA was 7.8% and 12.2% respectively. Bland Altman plots for the Cr2 splitting and [carnitine] are shown in Supplementary Fig. S3. Careful repositioning of the foot and leg within the coil helped minimise changes in splitting between visits, yet there was still some variability (Supplementary Fig. S3).

### The new method reveals carnitine associations with muscle mitochondrial oxidative capacity and lipid content

The 13 healthy volunteers (Dataset 1) were age-, BMI- and sex-matched to the 8 mitochondrial disease (MD) participants (Dataset 3) (mean ± SEM); age: 44.0 ± 3.9 vs 47.8 ± 5.3 years, p=0.57; BMI: 24.6 ± 0.8 vs 24.5 ± 1.6 kg/m^2^, p=0.87; sex: 6F,7M vs 3F,5M, p=0.72. One healthy volunteer withdrew from the study following acquisition of ^1^H-MRS of the VL, leaving n=12 in kPCr and ^1^H-MRS SOL and TA control measures. In one MD participant the TA measurement was not acquired (n=7 in TA), and in another the VL intramyocellular lipid (IMCL) and extramyocellular lipid (EMCL) resonances were unresolvable, so total lipid was included but individual IMCL, EMCL not.

#### Results in mitochondrial disease

Skeletal muscle *mitochondrial oxidative capacity* assessed by ^31^P-MRS was, as expected, significantly lower in participants with mitochondrial disease (MD) compared with healthy volunteers (1.90 ± 0.11 vs 2.23 ± 0.09 min^-1^; p = 0.036).

Skeletal muscle *total carnitine concentration* was lower in MD participants compared with healthy volunteers in both SOL (p=0.022) and VL (p=0.034), but not in TA (p=0.477) (Fig. 3A). There were no inter-muscle differences in total carnitine concentration in the healthy volunteers (all p > 0.88), but within the MD group the SOL and VL carnitine concentrations were lower than the TA (SOL p=0.011 and VL=0.012).

**Fig. 3.**
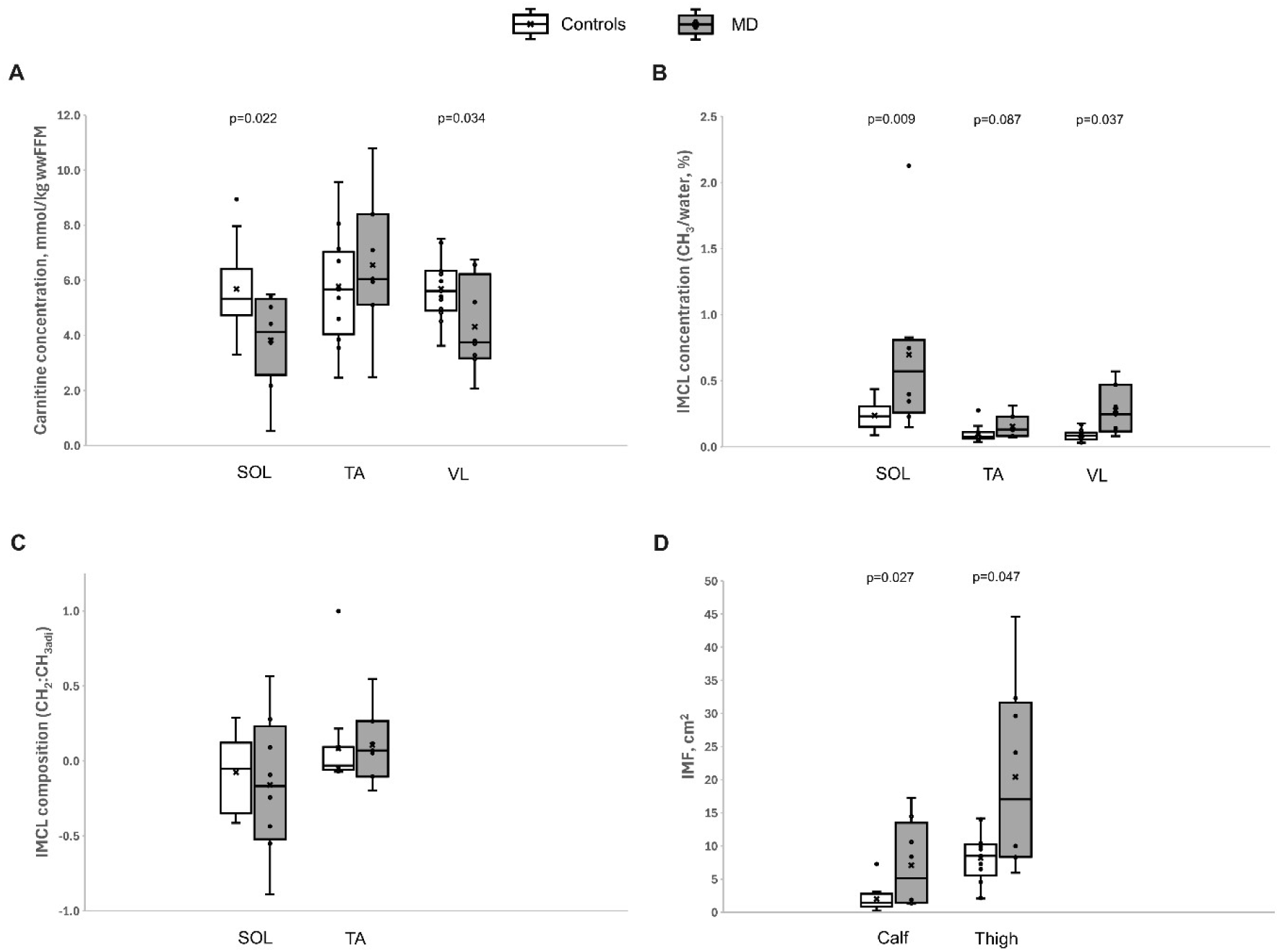
Carnitine concentrations and measures of lipid concentration and composition in mitochondrial disease (MD) patients and age- and BMI-matched controls. Soleus (SOL), tibialis anterior (TA) and vastus lateralis (VL) concentrations of carnitine (a) and intramyocellular lipid (IMCL). IMCL composition, assessed by the concentration adjusted CH_2_:CH_3_ ratio (c) and intramuscular lipid (IMF) assessed by magnetic resonance imaging (d), in controls (white bars) and patients with mitochondrial disease (grey bars).

Skeletal muscle *IMCL concentration*, as measured by the CH_3_ resonance, was higher in MD participants compared with healthy volunteers in SOL (p=0.009) and VL (p=0.037), with a similar tendency in TA which did not reach statistical significance (p=0.087) (Fig. 3B). *IMCL composition* (Fig. 3C) did not differ between the groups (both p>0.6).

*Intramuscular fat*, as measured as visible fat on T_1_-weighted imaging, which is dominated by TG in extramyocellular adipocytes, was higher in MD in both calf (p=0.027) and thigh (p=0.047) muscles (Fig. 3D).

There was no statistical difference in IMCL concentration or carnitine between the m.3243A>G/T and single deletion variants.

#### Local carnitine concentration is associated with local lipid content

Fig. 4A shows *relationships between carnitine and various lipid measures.* With all participants combined, the strongest relationships are within-muscle (so for example TA carnitine is unrelated to SOL or VL lipid contents) (Fig. 4A), suggesting between-muscle heterogeneity in both carnitine and lipid content (Fig. 4B). The strongest carnitine associations were with total lipid (EMCL+IMCL) (Fig. 4A). There were no significant associations with IMCL composition. Amounts in of EMCL (% of water, T_2_ corrected) were similar to IMCL within muscle groups in MD (SOL 1.6 vs 2.1, p=0.42; VL 1.9 vs 0.9, p=0.15) and healthy volunteers (SOL 0.9 vs 0.8, p=0.60), but slightly increased in TA and VL of MD (TA 0.7 vs 0.3, p=0.01, VL 0.7 vs 0.3, p=0.02) and TA of healthy volunteers (TA 1.9 vs 0.4, 0.01).

**Fig. 4.**
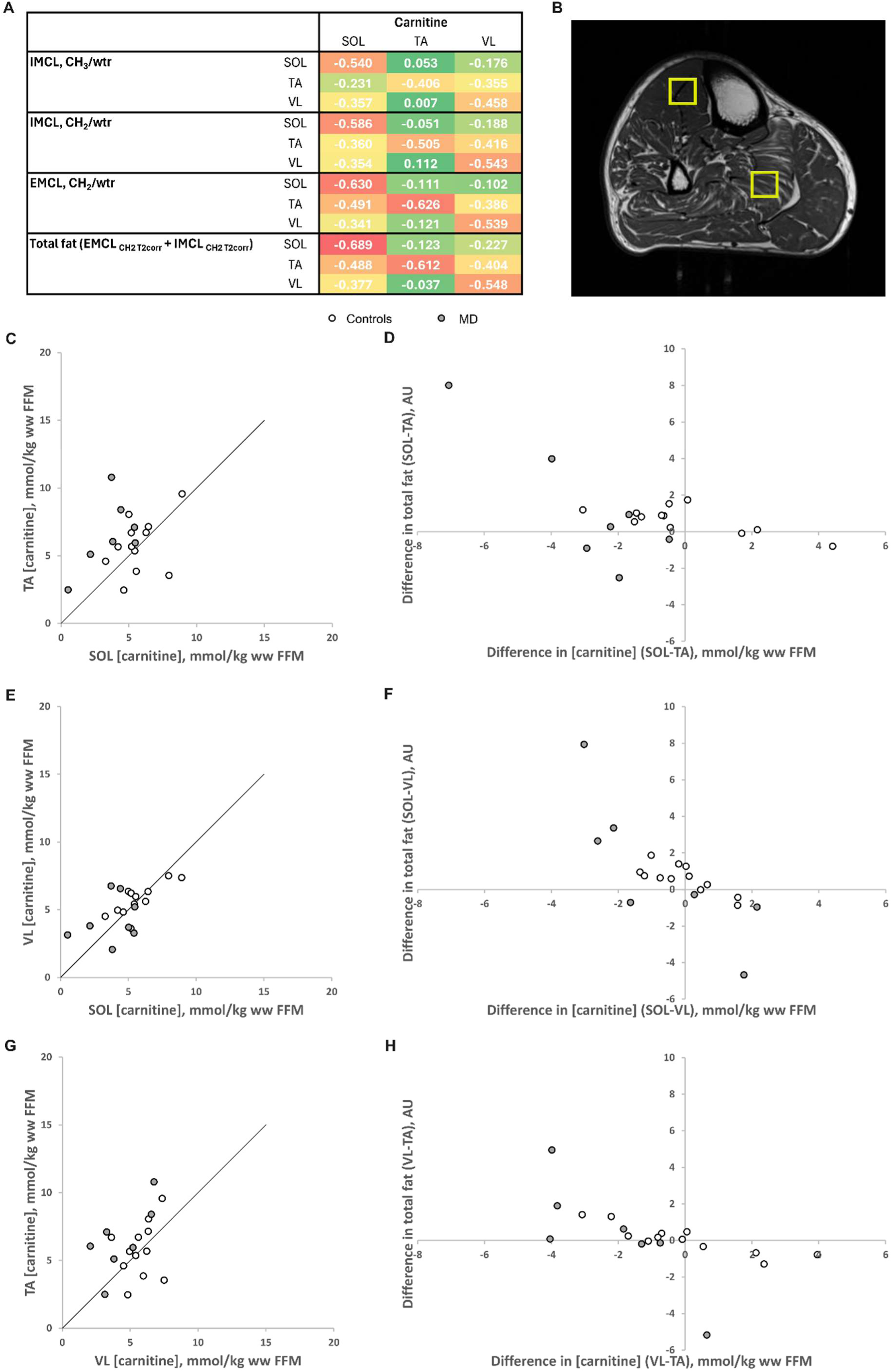
Relationship of [carnitine] with skeletal muscle lipid content. Table (a) shows Spearman’s correlation coefficients for soleus (SOL), tibialis anterior (TA) and vastus lateralis (VL) carnitine concentration and various lipid measures, with overlaid colours denoting strongest (red) to weakest (green) correlations. Image (b) gives representative voxel locations, placed to minimise visible fat. Note marked heterogeneity: carnitine concentration is 11.0 and 3.8 mmol/kg ww FFM for TA and SOL respectively. Graphs (c), (e) and (g) compare muscle carnitine concentration in TA *vs* SOL, VL *vs* SOL, and TA *vs* VL, respectively. Graphs (d), (f) and (h) compare the between-muscle differences in total fat (EMCL+IMCL) vs those in carnitine concentration between SOL and TA, SOL and VL, and VL and TA, respectively. Patients with mitochondrial disease and controls are denotes by grey and white circles respectively in (c-h).

Fig. 4C, E & G show the relationship of *carnitine concentration between muscles.* With all participants combined there was significant absolute agreement of [carnitine] between VL and SOL (ICC= 0.647, p=0.002), but not between TA and SOL or TA and VL (p=0.10 and p=0.07, respectively. The same pattern was seen in the healthy volunteers alone: significant absolute agreement between VL and SOL (ICC= 0.861, p=0.002), but not between TA and SOL or TA and VL (p=0.096 and p=0.231, respectively).

Figs. 4D, F & H show the relationship of *between-muscle variations in carnitine concentration with between-muscle variations in total lipid content* (EMCL+IMCL). For all participants together (n=19), the correlation was statistically significant for SOL *vs* VL (r_s_ = -0.782, p < 0.001) and VL *vs* TA (r_s_ = -0.786, p < 0.001) but not for SOL *vs* TA (r_s_ = -0.351, p = 0.137). Results were statistically significant in all comparisons in healthy volunteers alone (n=12): SOL *vs* VL, r_s_ = -0.779, p = 0.003; VL *vs* TA, r_s_ = -0.882, p < 0.001 and SOL vs TA, r_s_ = -0.733, p = 0.001.

The voxels were deliberately placed to avoid as much visible fat as possible in each muscle. The mean total fat, expressed as (EMCL_CH2 T2corr_ + IMCL_CH2 T2corr_) relative to the T_2_ corrected water signal, was for SOL, TA, and VL: (3.7, 2.3, 2.8 %) in MD and (1.7, 1.0, 1.1 %) in healthy volunteers. The highest total fat / water was 9% in MD, and 3% in healthy volunteers. The carnitine concentration is quantified relative to water, which we approximate to represent fat-free tissue. The small amounts of water present in adipose tissue cannot explain the range and correlations with carnitine we observe.

#### Mitochondrial oxidative capacity is more closely related to carnitine concentration than lipid content

The *relationship of muscle carnitine to mitochondrial oxidative capacity* is shown in Fig. 5A, which plots mitochondrial oxidative capacity (kPCr) measured in the quadriceps muscle with carnitine concentration in the VL (Fig. 5A): there is a good correlation in healthy volunteers alone (VL: r_s_ = 0.699, p = 0.001, n=12), while the relationship with mitochondrial disease participants does not reach statistical significance (VL: r_s_ = 0.397, p = 0.083, n=20).

**Fig. 5.**
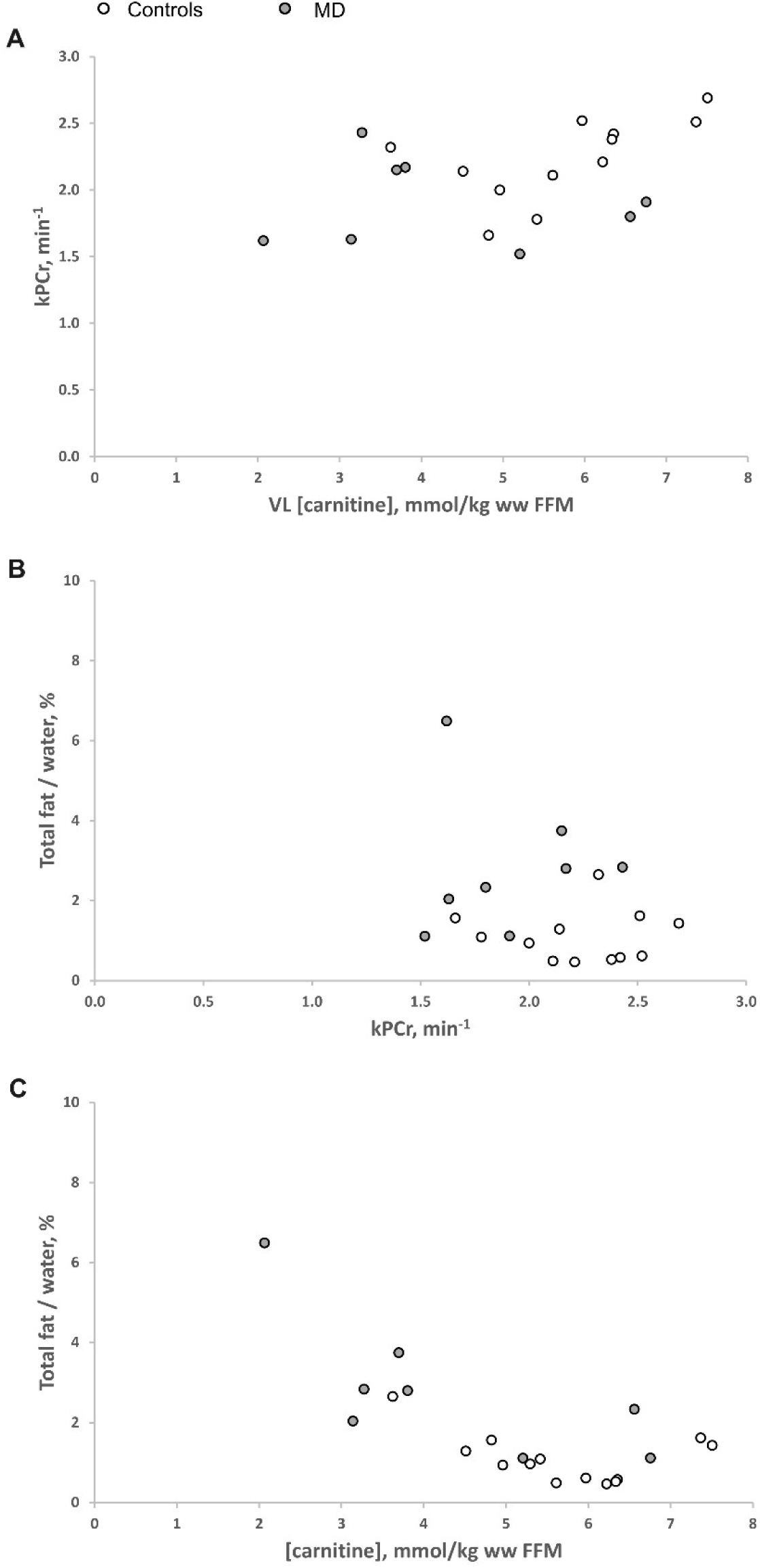
Relationship of mitochondrial oxidative capacity, carnitine concentration and lipid content in patients with mitochondrial disease, MD (grey circles) and healthy control participants (white circles). (a) mitochondrial oxidative capacity (kPCr) in the quadriceps *vs* vastus lateralis (VL) carnitine concentration. (b) kPCr *vs* VL total fat content (IMCL_CH2 T2corr_ + EMCL_CH2 T2corr_). (c) VL total fat (IMCL_CH2 T2corr_ + EMCL_CH2 T2corr_) content *vs* carnitine concentration

The *relationship of muscle lipid to mitochondrial oxidative capacity* is assessed in Fig. 5B. Quadriceps kPCr did not correlate statistically with any lipid measure in the VL (IMCL, EMCL, IMCL+EMCL; all p > 0.2) either in all participants combined or healthy volunteers alone.

VL carnitine concentration was associated with local (VL) total fat (Fig. 5C & 4A), as well as EMCL and IMCL (Fig. 4A).

#### Acetylcarnitine was not associated with mitochondrial oxidative capacity or lipid accumulation

*Acetylcarnitine* was measured in VL from a larger voxel of similar location to the carnitine measurement voxel. Its tendency to be increased in MD compared to healthy volunteers did not reach statistical significance (p=0.077; Fig. 6). Acetylcarnitine was not significantly correlated with kPCr either in all participants combined or healthy volunteers alone (both p > 0.4), nor was it associated with any lipid measure from the small voxel (all p > 0.4) or total lipid from the larger acetylcarnitine voxel (both p > 0.2).

**Fig. 6.**
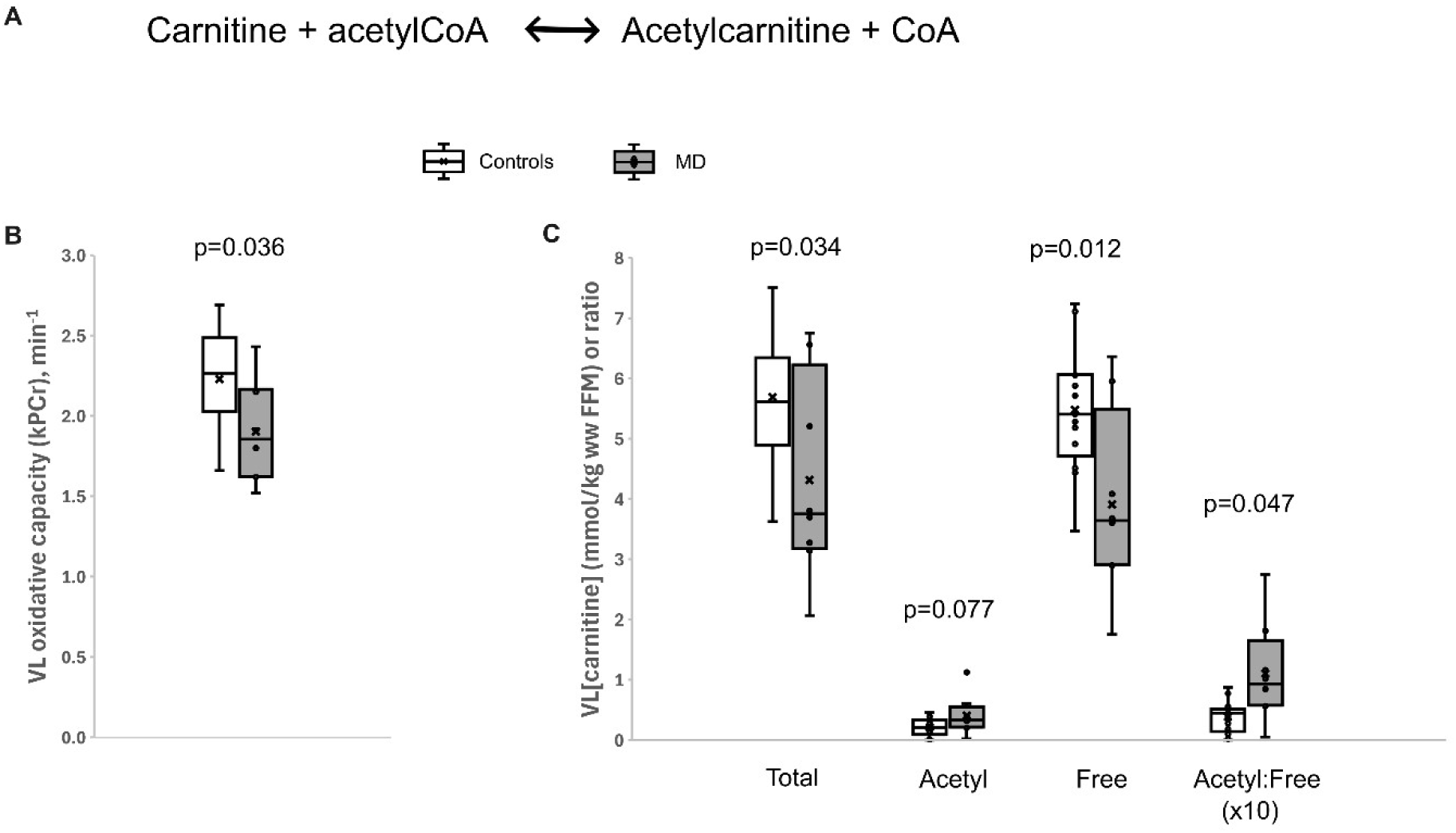
Mitochondrial oxidative capacity and different carnitine measures in mitochondrial disease (MD) and healthy control participants. (a) Relation between carnitine and acetylcarnitine within the mitochondria, facilitated by CrAT and used as a coping mechanism to buffer excess acetyl CoA in situations such as nutrient excess or mitochondrial dysfunction. (b) Mitochondrial oxidative capacity in MD (grey boxes) and controls (white boxes). (c) MR measures of total carnitine, acetylcarnitine, ‘free’ carnitine, and acetyl:free carnitine, in controls (white boxes) and MD (grey boxes).

*Free carnitine* was estimated approximately (as voxel locations differ and this does not consider medium and long chain acylcarnitines), as acetylcarnitine subtracted from total carnitine. It was significantly lower in MD (p=0.012; Fig. 6). In all participants combined free carnitine showed similar and stronger correlations to kPCr as total carnitine (r_s_ = 0.453, p=0.045 versus r_s_ = 0.397, p = 0.083). In the healthy volunteers alone, similar associations were found between kPCr and either free or total carnitine. The *acetylation status*, defined as the ratio of acetylcarnitine: ‘free’ carnitine, was significantly raised in the MD participants (p=0.047; Fig. 6).

## Discussion

We have described and validated a novel method for quantifying skeletal muscle total carnitine concentrations that uses intrinsic features of the ^1^H spectrum to correct for fibre orientation and to achieve high analytical specificity relative to other metabolites contributing to the TMA region. The orientation-dependent visibility correction, quantified from the splitting of the creatine methylene peak is, as expected, significant between muscles but importantly we have shown that it is also necessary even within a single muscle. For example, in soleus which is typically thought to be close to the magic angle (where theoretically no orientation correction would be necessary), this effect costs between 0 % and 40 % of signal despite standardised leg positioning.

We have also demonstrated that the TMA resonance largely reflects total carnitine concentration (as previously suggested ^11–13^), but likely includes a smaller carnosine component (amounting to an equivalent TMA concentration of ∼ 1.3 mmol/kg ww), for which we corrected using the downfield carnosine resonance. We did not find taurine to be a significant contributor of the TMA resonance, although of course this will need to be checked in other disease states. Our method yields human skeletal muscle total carnitine concentrations in agreement with biopsy measurements in the same individuals, and mean healthy volunteer carnitine concentrations (∼ 6 mmol/kg ww) that are well within the expected range of 3-8 mmol/kg ww ^18–20^. Although we have demonstrated this at 3T, this methodology, adapted for field-strength relevant relaxation and invisibility phenomenon, would also work at higher field strengths where it may be easier to implement due to increased SNR and reduced influence of creatine splitting in the TMA region.

We found that in patients with mitochondrial disease compared to controls, total carnitine was lower in the soleus and vastus lateralis, although not significantly so in tibialis anterior. This may suggest some fibre-type dependence, the soleus and vastus being more oxidative muscles and more reliant on fat oxidation, which may be more likely to accumulate acylcarnitine’s and export them, thereby reducing the total carnitine concentration. This theory is consistent with previous findings ^21^, where aging-associated metabolic block in the TCA cycle was associated with reduced carnitine, predominantly in the red soleus muscle. It could also be influenced by a lack of statistical power, as the power calculations were based on soleus IMCL levels, and quadriceps measures of mitochondrial function.

In the opposite direction to tissue carnitine, the IMCL concentration in the mitochondrial disease patients compared with the healthy volunteers was higher in soleus and vastus lateralis (and tendency in tibialis anterior). This association of lowered carnitine and raised lipid can also be seen in the significant correlations of total carnitine with same-muscle IMCL, EMCL, and IMCL+EMCL; by contrast, carnitine concentrations were only weakly associated if at all with lipid in other muscles, demonstrating the spatial heterogeneity in both carnitine and lipid within a single individual. Indeed, this was further supported by significant correlations of between-muscle differences in total carnitine with differences in total lipids, which remained significant in healthy volunteers alone, suggesting this mechanism is operative in health as well as disease. A strong inverse association of carnitine to lipid concentration has been previously demonstrated in biopsy samples of rat red muscle ^22^. It is important to note that unlike free and total carnitine, acetylcarnitine had a tendency to be increased in mitochondrial disease and was not related to any measures of lipid or mitochondrial oxidative capacity.

Thus, mitochondrial disease is associated with reduced skeletal muscle total carnitine concentration and raised muscle lipid in the more oxidative soleus and vastus lateralis muscle, with no alteration in lipid composition. As no mitochondrial genes are known to alter carnitine synthesis or uptake, this suggests that mitochondrial dysfunction can cause localised carnitine depletion due to increased export of acylcarnitines. It is unclear whether the lipid accumulates as a direct result of reduced fatty acid oxidation, or if carnitine governs the rate of fatty acid oxidation. It is interesting to speculate whether this carnitine-lipid association may be indicative of heterogeneity in mitochondrial function, and if so, whether this extends to a cellular level, which would be consistent with highly localised brain effects in mitochondrial diseases such as MELAS, and the spatial heterogeneity of heteroplasmy ^23^.

As this association of mitochondrial oxidative capacity and carnitine as well as spatial differences in carnitine and lipid exists within the healthy volunteers, we speculate that this mechanism may underlie aging-associated mitochondrial dysfunction and skeletal muscle lipid accumulation, and associated muscle fatigue / sarcopenia. As soleus IMCL concentration (unlike IMCL composition) is also associated with total body fat ^24^, it may also be that this metabolic block originating from intrinsic mitochondrial dysfunction and/or decreased carnitine could, in the long term, contribute to the pathogenesis of obesity.

Of course, the reverse may occur where obesity leads to lipid oversupply to muscle, exceeding mitochondrial demand and leading to increased acylcarnitine export, resulting in reduced total carnitine content and a resultant metabolic block. This has been demonstrated by Noland et al ^25^, where rats fed a lifelong high fat diet, had compromised carnitine levels that corresponded with increased skeletal muscle accumulation of acylcarnitine esters and diminished hepatic expression of carnitine biosynthetic genes. However, this is unlikely the only mechanism in health as it does not easily explain the intrasubject lipid and carnitine heterogeneity that is observed.

Another possibility is that low skeletal muscle carnitine results from reduced cellular uptake. Experimental inhibition of carnitine biosynthesis in rats was shown to cause impaired exercise tolerance, muscle atrophy, impaired electron transport chain function, and increased free radical leak in the soleus muscle ^26^. However, this alone could not easily explain the observed intrasubject heterogeneity, and so although each mechanism may contribute to carnitine concentration, the intrasubject heterogeneity is most likely to originate from heterogeneity in mitochondrial dysfunction.

In any case, carnitine supplementation has been shown to ameliorate both mitochondrial impairment and lipid accumulation in mitochondrial disease ^2,27,28^. A study demonstrating that reduced fatty acid oxidation due to low carnitine levels was reversed to control levels with addition of carnitine in a patient’s muscle homogenates ^29^, and evidence that carnitine deficient larvae die ‘fat’ when starved and unable to use their fat stores ^30^, does imply that carnitine can influence the ability to oxidise fat. A meta-analysis of randomised controlled trials in humans has demonstrated significant benefits of carnitine supplementation on weight, BMI, and fat-mass ^4^.

### Potential clinical utility of the carnitine measurement

As noted within the Introduction, plasma carnitine concentration is not representative of tissue concentrations, and cannot help in assessing whether a carnitine supplement is reaching its target tissue. Our novel non-invasive method for measuring skeletal muscle total and free carnitine concentration could help with this, given its non-invasive nature, reproducibility, and ability to perform longitudinal studies on the same sample of tissue. Although the carnitine-deficient may have most to gain from therapeutic carnitine supplementation, a repeatable and reliable measurement, such as we have described here, should be valuable in exploring the potential relevance and benefit of carnitine to human health.

## Methods

### Participants and preparation

#### Dataset 1: Healthy volunteer dataset used in a) method validation and b) age- and BMI-matched controls to Dataset 3

13 healthy volunteers (6F, 7M) were recruited through advertisement. Exclusion criteria were current smoking, drug or alcohol addiction, any relevant medical disorder (e.g. Type II diabetes) or medication (e.g. statins) that could influence measurements, and standard MRI contraindications. Volunteers maintained a normal diet for 3 days and refrained from alcohol and vigorous activity for at least 19 h prior. A light breakfast of toast or cereal was served at the National Institute for Health and Care Research (NIHR) Cambridge Clinical Research Facility (CRF) prior to ^1^H MRS.

#### Dataset 2: Method reproducibility in healthy subjects

An existing dataset of 24 healthy non-obese Caucasian male volunteers, aged 18 to 50 years without diabetes and taking no medication ^31,32^. They were asked to refrain from alcohol and vigorous physical activity and to follow their normal diet for 3 days prior to the study. The study took place at the NIHR Cambridge CRF and their meal on the evening prior to the study (at 19:30) and following breakfast on day 1 (at 07:30) were standardized based on one-third of the recommended daily intake of energy and contained approximately 50% carbohydrate, 30% fat, and 20% protein. Subjects fasted from 08:00 on day 1 to 14:00 on day 2. MRS was undertaken at 8 and 28 h of fasting (16:00 on day 1 and noon on day 2), with the voxels being carefully relocalised on the second visit. Spectra with no clear distinction between EMCL and IMCL CH_2_ resonances were eliminated, yielding 22 and 19 complete pre- and post-fasting sets for TA and SOL respectively ^32^.

#### Dataset 3: Application to mitochondrial disease

8 participants with mitochondrial disease due to a genetically confirmed heteroplasmic mitochondrial DNA disorder were recruited from the Cambridge mitochondrial clinic (4 with m.3243A>G, 1 with m.3243A>T, and 3 with single large mtDNA deletion (CPEO)). Exclusion criteria were clinically significant liver disease or abnormal liver function tests, random plasma glucose > 20mMol/L in screening bloods, presence of significant other neurological disorders or major co-morbidities (such as definite cognitive impairment, psychiatric disease, heart, or lung failure, orthopaedic or rheumatological disorders), MRI contraindications, and females taking the combined oral contraceptive pill.

The East of England Cambridge Central Ethics Committee (REC 06/Q0108/84), Cambridge Local Research Ethics Committee, and East of England Cambridge South Research Ethics Committee, approved the healthy volunteer reference measurement, fasting healthy volunteer, and patient studies respectively.

### Measurements of total carnitine concentration

Our novel ^1^H MRS method comprises three parts. It uses optimised fitting incorporating prior knowledge of the Cr3 triplet frequencies, applies an NMR visibility correction dependent based on the Cr2 splitting, and allows for other metabolites that may resonate at the TMA frequencies. This is summarised in Supplementary Figure 4.

### ^1^H-MRS acquisition

All acquisitions used a Siemens Magnetom 3T Verio or Skyra_fit_ (Erlangen, Germany) with the peripheral-angio (PA) coil. The participants were placed supine, legs on the Siemens PA leg-rest, and with feet pointing towards the ceiling (PA coil and leg-rest placed to enable PA struts to enforce this, which provides consistency in participant positioning as well as reducing likelihood of movement). ^1^H MRS was acquired with a short echo time of 35 ms, using the point-resolved spectroscopy sequence. A water-suppressed spectrum was acquired from a voxel of dimension 13 mm, positioned to avoid as much visible fat on T_1_-weighted images within the soleus (SOL), tibialis anterior (TA), and vastus lateralis (VL) (Datasets 1 & 3) muscles as possible. Using a 5 second repetition time, 32 averages (or 64 averages in dataset 2) of water-suppressed data were acquired, and a non-water suppressed spectra also acquired in Datasets 1 & 3.

### Optimised fitting

Data were analysed in jMRUI ^33,34^ and fitted with the AMARES algorithm ^35^ using two fitting files modified from Savage et al. ^24,36^. Fitting routine 1 additionally included a singlet for carnosine C2, as well as a doublet for the total creatine CH_2_ resonances (Supplementary Table 1) and was used to measure all lipids, carnosine as well as the frequency splitting of the creatine CH_2_ peaks. In cases where the fitting failed to correctly fit the Cr2 resonances, the splitting was measured manually. Fitting routine 2 included a triplet for the total creatine CH_3_ resonances, which encompassed inferred prior knowledge of the CH_3_ frequencies relative to CH_2_ (Supplementary Table 1). We found that this frequency prior knowledge permitted some soft constraint in the actual frequency (hence the necessity to run a two-step fitting approach), and this fit was used to obtain measures of TMA and creatine CH_3_.

### Correcting for orientation-dependent NMR visibility signal loss

Both the TMA and Cr3 resonances exhibit NMR visibility effects: when the muscle fibres are parallel to the main magnetic field, the resonances are 55% and 29% lower, respectively, than at the magic angle at 3T ^14^. The angle of the fibres relative to the main magnetic field can be conveniently calculated by fitting the frequency splitting of the total creatine (Cr2) resonance at ∼3.95 ppm. Fig. 1B illustrates the Cr2 splitting (x, in Hz) and hypothetical signal loss of TMA and Cr3 assuming a linear signal decrease with Cr2 splitting (x) ^14^. In order to correct the signal (S), of TMA or summed Cr3 (Cr3+Cr3a+Cr3b), for its associated orientation-dependent loss we used our splitting values for the average parallel angle (tibialis anterior, 20.5 Hz) and magic angle (we measured ∼4 Hz was still a singlet), to generate an equation for the orientation-corrected signal (S_corr_), such that,

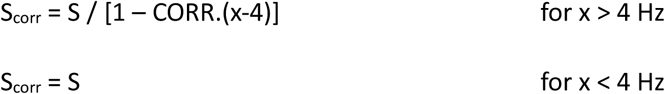

where x is the frequency difference (Hz) between the two Cr2 resonances, and CORR is a scaling factor of 0.0333 and 0.0176 for TMA and summed Cr3 respectively.

### Correcting for orientation-dependent T2 relaxation times

Water T2 relaxation time is mildly orientation dependent ^14^ and hence using our magic (4 Hz) and parallel (20.5 Hz) angles we established:

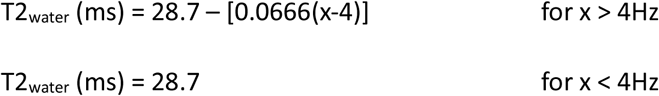

Given that the TMA T2 is likely intertwined with NMR visibility, we applied the NMR visibility correction and used a constant T2 of 134 ms ^13^. Established orientation-independent T2 values of creatine were used (135, 135 and 162 ms for the soleus, tibialis anterior, and vastus lateralis muscles respectively)^10^, with an additional fixed factor assuming a short decay component plateau to 2/3 ^37,38^.

The T2 relaxation time of the C2 carnosine resonance at ∼8 ppm has been shown to be strongly orientation-dependent ^39^. Using the T2 values within this and converting from residual dipolar coupling (D) to peak-to-peak splitting (x) (personal correspondence), it was found that the T2 (ms) of carnosine-C2 (T2 _C2_) would be:

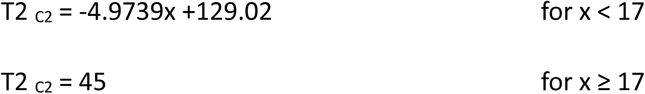

where x is the frequency Cr2 splitting (Hz). An upper limit of 17 Hz was set as this was their upper measurement limit, and to also prevent amplified errors due to the exponential nature of T2 correction.

### Validation of NMR visibility correction

Many MRS quantitation methods depend on the fact that water and creatine concentrations in human skeletal muscle vary little between subjects or muscles. As the Cr3 resonance exhibits orientation-dependent NMR invisibility effects, we were able to test the performance of our orientation-dependent correction on the Cr3 resonance by comparing the water – creatine calibration conversion.

Using Dataset 1, a peak within the spectrum (TMA) was compared relative to water and also relative to the summed Cr3 triplet. The TMA_corr_, water, and sumCr3_corr_ were corrected for T_2_ relaxation effects as outlined above, apart from in the case of ‘without NMR visibility correction’ where the water T2 was 28 ms to simulate the case where no orientation-dependent correction is undertaken. With the Cr3 triplet uncorrected (Fig 1C) large deviations are found between muscle groups and calculated Cr levels outside expected levels within the TA. Upon orientation-dependent visibility correction of the Cr3 triplet (Fig 1D) this appears corrected. In addition to informing on the effectiveness of the NMR visibility correction, this also provides calibration for the conversion of datasets that did not acquire non-water suppressed data (Dataset 2).

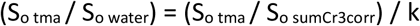

where S_o_ is the NMR visibility and T2 corrected signal intensity of the resonance, and k is the calibration conversion factor (see results section).

### Calculation of absolute concentrations and consideration of other metabolites

Absolute concentrations of carnitine (CTN) in mmol/kg wet weight fat free mass (mmol/kg ww FFM) were calculated with standard assumptions regarding muscle water content and proton density:

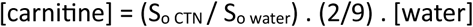

Where S_o_ is the NMR visibility and T2 corrected signal intensity of the resonance, [water] is the concentration of water in skeletal muscle (calculated using a pure water concentration of 55,342 mmol/l and assuming a relative tissue water content of 0.804 kg/kg ^40^ and tissue density of 1.05 g/ml^41^).

Other metabolites which resonate at similar frequencies to carnitine could contribute to the TMA peak (Table 1). However, only free carnitine, acetyl and acyl-carnitines, and carnosine are quantitatively important (see Results). Carnosine is most reliably quantified from the C2 resonance at ∼8 ppm. Assuming the carnosine resonance within the TMA region has the same NMR invisibility and T2 as the carnitine which makes up most of the TMA pool, carnitine (CTN) levels can be quantified as:

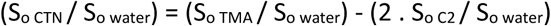

where S_o_ is the visibility- and T2-corrected signal intensity of the resonance, and C2 is carnosine at ∼8ppm.

In order to establish if taurine is significantly contributing to the TMA signal, we investigated the downfield resonance (∼3.4ppm) relative to expected signal here from free carnitine alone. The orientation-dependent visibility and T2 corrected signal of this downfield resonance (S_o 3.4_) was calculated assuming the same T2 as the TMA resonance, and corrected for NMR visibility using signal loss of 67 % at the parallel angle ^14^. The ∼3.4 ppm resonance was fitted by 2 gaussian peaks with soft constraints on frequencies: 3.33-3.40 ppm and 3.40-3.45 ppm. Linewidth of one peak had soft constraints of 5-10 Hz, while the second peak equal to that of the first.

### Validation against skeletal muscle biopsy

Estimated carnitine concentrations (mmol/kg ww FFM) were validated against total carnitine concentrations (mmol/kg ww) measured in biopsy samples taken in 4 participants within dataset 3.

### Muscle biopsy and analysis

Assay of free and acyl carnitines in the tissue samples used a pre-analytical chemical derivatization - UPLC-MS/MS method with 3-nitrophenylhydrazine as the derivatizing reagent^42^. A stock solution containing authentic standards of all measured carnitines was prepared in 85% HPLC-grade ethanol. The stock solution was serially diluted with the same solvent to prepare ten calibration solutions. Concentrations of each carnitine in the calibration solutions ranged from 0.01 nM to 20 µM. Next, for sample preparation, each tissue sample was precisely weighed in a 1.5-mL safe-lock Eppendorf tube. 1.5 μL of water per mg of raw tissue was added to the tube. The tissue was homogenized using two 3-mm metal beads on an MM 400 mill mixer at 30 Hz for 2 min. After 3-s spin-down on a microcentrifuge, 8.5 μL of HPL-grade ethanol per mg of raw tissue was added. The tissue was homogenized again for 3 min, then centrifuged at 21,000 g at 5 °C for 10 min.

20 μL of the clear supernatant from each sample or 20 μL of each calibration solution was mixed in turn with 20 μL of an internal standard solution containing 20 deuterium-labeled free and acyl carnitines, 40 μL of a 150-mM 3-nitrophenylhydrazine. HCl solution in 60% methanol and 40 μL of a 120 mM EDC.HCl-5% pyridine solution in 60% methanol. The mixtures were incubated at 40 °C for 30 min. After reaction, 10 μL aliquots of the resulting calibration and sample solutions were injected into a Waters BEH C18 column (1.7 µm, 2.1 mm ID, 10-cm long) for the UPLC-MS/MS analysis on an Agilent 1290 Infinity II UHPLC coupled to an Agilent 6495C triple-quadrupole mass spectrometer. The mass spectrometer was operated in the positive-ion electrospray ionization mode. A binary-solvent mobile phase composed of 0.1% formic acid in water and 0.1% formic acid in acetonitrile-isopropanol (1:1) was used for gradient elution under optimized chromatographic separation and MS/MS detection (multiple reaction monitoring). The UPLC-MS/MS data files were recorded and processed using Agilent *MassHunter*® 10.0. Linear regression calibration curves for individual carnitines were constructed using data acquired from the calibration solutions. Concentrations of carnitines in the samples were calculated by interpolating the calibration curves using data acquired from the sample solutions, within an appropriate concentration range for each compound.

### Reproducibility study

An existing dataset (Dataset 2) ^31,32^ was used to assess reproducibility of the carnitine measurement. As this included an intervention of a fast, with unknown effects on skeletal muscle carnitine concentrations, this measure represents an upper limit of reproducibility.

### Application of carnitine measure to mitochondrial disease and lipid accumulation

#### Hypothesis and power calculations

We hypothesised that mitochondrial dysfunction would be associated with carnitine depletion and lipid accumulation, and tested this by comparing patients with primary mitochondrial disease with age-, sex- and BMI-matched healthy volunteers.

In the absence of published data on muscle carnitine concentrations we based our power calculations on the lipid (IMCL) endpoint and mitochondrial oxidative capacity (kPCr) measures, assuming this would provide sufficient power for carnitine. <u>IMCL concentrations</u> were estimated previously in MELAS to be 26 ± 18 mmol/kg ww ^11^, and in healthy volunteers 7.8 ± 3.5 mmol/kg ww ^36^, both in soleus muscle. Using 90 % power, 0.7 ratio enrolment, and pooled standard deviation yields n of 4 and 5 for mitochondrial disease and healthy volunteers respectively. <u>Mitochondrial oxidative capacity</u>: Using measures reported within ^43^, 90 % power, 0.7 ratio enrolment, and pooled standard deviation yields n of 5 and 7 for mitochondrial disease and healthy volunteers respectively.

### Methods

13 age-, BMI-, and sex-matched healthy volunteers (Dataset 1) were matched to 8 participants with mitochondrial disease (Dataset 3) and underwent ^1^H-MRS and ^31^P-MRS assessments of [carnitine], lipids, and mitochondrial oxidative capacity, respectively, on a 3T Siemens Skyra_fit_.

### ^1^H-MRS and MRI

Acquired in soleus, tibialis anterior, and vastus lateralis muscles both with and without water suppression. IMCL concentration was expressed as a percentage of IMCL CH_3_/water. The IMCL compositional marker, CH_2_CH_3adj_ was calculated as described previously ^24^. Intramuscular fat (IMF) was quantified from T1-weighted images (MRI) as outlined before ^32^. Single voxel spectroscopy total fat content was represented by the sum of T_2_-corrected EMCL CH_2_ and IMCL CH_2_ relative to water, where T2 values used were EMCL 77.6/77.5 ms, IMCL 89.4/90.9 ms for the soleus/tibialis anterior muscles respectively ^44^, and where water T2 was calculated as described above.

### Acetylcarnitine

Acquired in vastus lateralis/intermedius from a voxel 40x25x40 mm. The acetylcarnitine resonance at 2.1 ppm was measured at long echo time. Acetylcarnitine and water were measured with echo times 500 and 35 ms and corrected for T2 relaxation effects using T2 values of 265 and 28 ms, respectively. Absolute concentrations were calculated by correcting for proton density (2/3) and accounting for the concentration of water in skeletal muscle as outlined above.

### ^31^P-MRS

Mitochondrial oxidative capacity was assessed by ^31^P magnetic resonance spectroscopy measure of the rate constant of phosphocreatine resynthesis post exercise using established methods ^45–47^, comprising a standardised exercise protocol of two bouts of knee extensions, each 1 minute long at 0.5 Hz, separated by 5 minutes of rest.

### Statistics

Statistical analysis was performed in SPSS Statistics 28 (IBM, Armonk, NY, USA), setting significance at p < 0.05. Data that was not normally distributed (by Shapiro–Wilk test) was logarithmically transformed prior to statistical analysis. Linear regression analysis was used to compare signals relative to creatine and water in each muscle group. Absolute agreement of MRS carnitine measures with biopsy were assessed by the intraclass correlation coefficient (ICC) for single measures. Reproducibility was assessed by Coefficient of Variation and Bland Altman analysis.

Two-tailed independent samples t-tests was used to test for group differences between datasets 1 & 3. In the exceptional case where a non-normally distributed variable could not be logarithmically transformed due to negative values (TA IMCL CH_2_CH_3adj_), the Mann U-Whitney test was performed. A paired-sampled t-test was used to test for differences in carnitine concentration between muscles. Spearman’s correlation coefficient was used to test the associations as some relations were non-linear and to also account for outliers. Absolute agreement of carnitine concentration between muscle groups was assessed by the intraclass correlation coefficient (ICC).

## Data availability

Datasets from participants who consented to data sharing are available from the corresponding author upon reasonable request.

## Acknowledgements

We thank all the participants, the staff at the NIHR Cambridge Clinical Research Facility and Wolfson Brain Imaging Centre, as well as Sarah Nutland (NIHR Cambridge BioResource). We acknowledge the UVic-Genome BC Proteomics Centre, Victoria, Canada for targeted metabolomic analysis. This work was supported by the National Institute for Health and Care Research (NIHR) Cambridge Clinical Research Facility and the NIHR Cambridge Biomedical Research Centre [BRC 1215 20014, NIHR203312], Medical Research Council [MR/V011758/1], the Wellcome Trust [210755/Z/18/Z, 212219/Z/18/Z], Addenbrooke’s Charitable Trust, and the British Society for Pediatric Endocrinology and Diabetes. KRS was an Academic Clinical Lecturer funded by the UK NIHR and she was supported by MRC International Centre for Genomic Medicine in Neuromuscular Disease (ICGNMD) [MR/S005021/1] and is supported by the LifeArc Centre to Treat Mitochondrial Diseases (LAC-TreatMito). JvdA is supported by a Wellcome Clinical Research Career Development Fellowship (219615/Z/19/Z) and a Wellcome Discovery Award (226653/Z/22/Z). R.H. is supported by the Wellcome Discovery Award [226653/Z/22/Z], the Medical Research Council (UK) [MR/V009346/1], the Hereditary Neuropathy Foundation, AFM-Telethon, Ataxia UK, Action for AT, the A-T Children’s Project, Muscular Dystrophy UK, CureARS, the Rosetrees Trust [PGL23/100048], the LifeArc Centre to Treat Mitochondrial Diseases (LAC-TreatMito) and the UKRI/Horizon Europe Guarantee MSCA Doctoral Network Programme (Project 101120256: MMM). She is also supported by an MRC strategic award to establish an International Centre for Genomic Medicine in Neuromuscular Diseases [MR/S005021/1]. PFC is currently funded by a Wellcome Discovery Award [226653/Z/22/Z], a Wellcome Collaborative Award [224486/Z/21/Z], the Medical Research Council Mitochondrial Biology Unit [MC_UU_00028/7], and the Biotechnology and Biological Sciences Research Council [BB/Y003209/1], the Rosetrees Trust [PGL23/100048], and the LifeArc Centre to Treat Mitochondrial Diseases (LAC-TreatMito) under grant no. 10748. LifeArc is a charity registered in England and Wales under no. 1015243 and in Scotland under no. SC037861. AS is supported by the NIHR Cambridge Clinical Research Facility. This is a summary of independent research funded by the Medical Research Council and National Institute for Health and Care Research (NIHR) and carried out at the NIHR Cambridge Clinical Research Facility (CRF). The views expressed are those of the authors and not necessarily those of the NIHR or the Department of Health and Social Care.

