## Supplementary for "Novel *in vivo* measurement of muscle total carnitine concentration reveals potential mechanism linking mitochondrial dysfunction and lipid accumulation"

### Supplementary information

**Table S1. Fitting routines and parameters.**

| | Fitting routine 2 (TMA, Cr3 and Cr3a,b) | | | | | | | | | |
|---|---|---|---|---|---|---|---|---|---|---|
| | Fitting routine 1 (Carn-C2, Cr2a,b Hz and EMCL, IMCL $CH_2$ and $CH_3$) | | | | | | | | | |
| | Resonance | | | | | | | | | |
| | **Carn-C2** | **Water** | **Cr2a** | **Cr2b** | **TMA** | **Cr3** | **EMCL $CH_2$ (E2)** | **IMCL $CH_2$ (I2)** | **E/IMCL $CH_3$ (E3, I3)** | **Cr3a,b** |
| **Starting values** | | | | | | | | | | |
| Frequency, ppm | 7.99 | 4.7 | 3.97 | 3.85 | 3.2 | 3.02 | 1.5, | 1.29 | 1.09, 0.9 | 3.07, 2.94 |
| **Prior knowledge** | | | | | | | | | | |
| Frequency, ppm | 7.91-8.03 | 4.4-5.0 | 3.9-4.05 | 3.75-3.9 | 3.16-3.22 | 2.95-3.16 | 1.46-1.54 | 1.21-1.35 | E2 – c, I2 - c | Cr2a - k Cr2b- k’ |
| Linewidth, Hz | 3-15 | Free | 0-15 | 0-15 | 0-15 | 0-15 | 0-30 | 0-30 | E2 + α, I2 + α | Cr3 |
| Relative phase | 0 | 0 | $0\pm30^0$ | $0\pm30^0$ | 0 | 0 | 0 | 0 | 0 | 0 |
| Resonance area | Free | Free | Free | Cr2a | Free | Free | Free | Free | Free | Free |
| Lineshape | Gau | Lor | Gau | Gau | Gau | Gau | Gau | Gau | Gau | Gau |
| **Overall Phases** | | | | | | | | | | |
| Phases and weighting | PH0=0; PH1=0; Weighting points 1-5; Truncated points=0 | | | | | | | | | |

Carn-C2, C2 carnosine resonance around 8 ppm; Cr2a, total creatine $CH_2$ downfield resonance; Cr2b, total creatine $CH_2$ upfield resonance; TMA, trimethylammonium compounds; Cr3, total creatine $CH_3$ main resonance; Cr3a, creatine $CH_3$ downfield resonance; Cr3b, creatine $CH_3$ upfield resonance; EMCL, extramyocellular lipid; IMCL, intramyocellular lipid; $CH_2$, methylene protons; $CH_3$, methyl protons. E2, EMCL $CH_2$; I2, IMCL $CH_2$. α, a value of approximately 2.5 Hz; c, a constant of 50.4 Hz; k, a constant of 110 Hz; k’, a constant of 110 Hz except TA where is 113 Hz. PH0, zero order phase; PH1, first order phase (begin time).

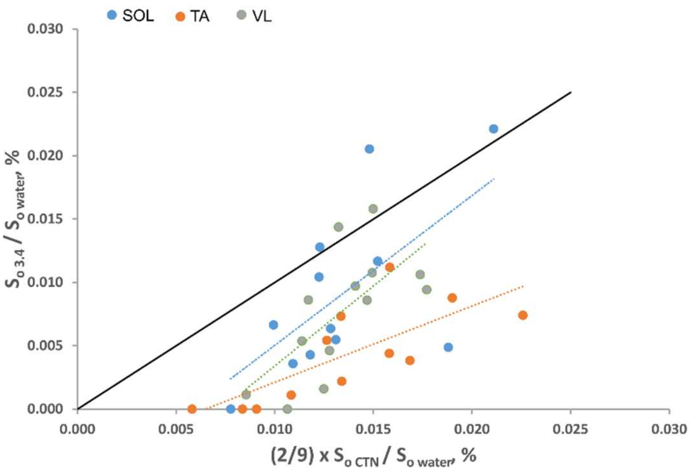


**Fig. S2.** Signal ($S_o$) corrected for orientation-visibility and T2 at 3.4 ppm vs expected carnitine signal at this frequency in healthy volunteers (soleus in blue, vastus lateralis in green, and tibialis anterior in orange). Solid line is the line of identity; same-colour dotted lines are linear regression lines for each muscle.

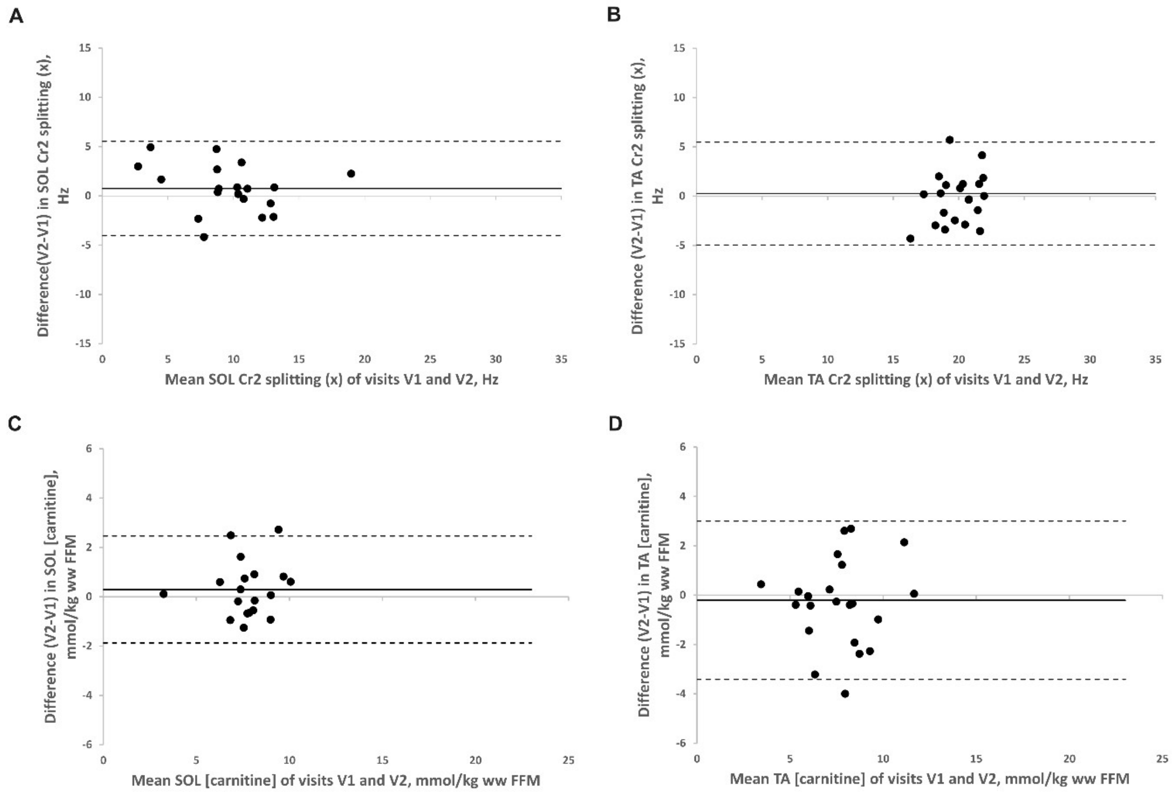


**Fig. S3. Bland Altman plots** for repeatability of Cr2 splittings (a,b) and carnitine concentrations (c,d), in soleus (a,c) and tibialis anterior (b,d). The mean difference is denoted by the solid horizontal line and the limits of agreement (±1.96 x SD) from the mean difference by the dotted lines.

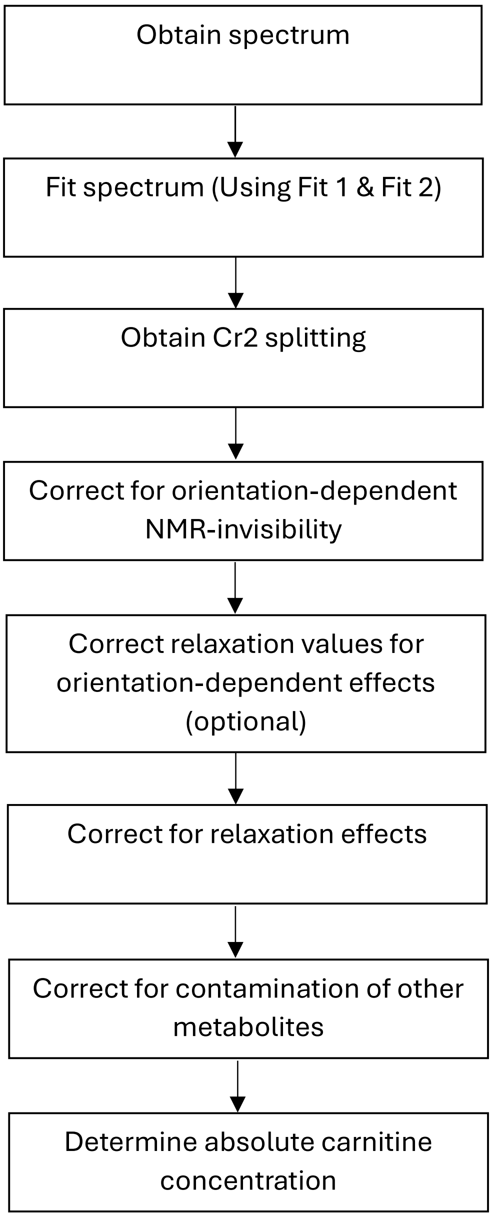


**Fig. S4. Flow chart summary detailing the methodology**
